# Proteomic and metabolomic signatures associated with the immune response in healthy individuals immunized with an inactivated SARS-CoV-2 vaccine

**DOI:** 10.1101/2021.07.21.21260959

**Authors:** Yi Wang, Xiaoxia Wang, Laurence Don Wai Luu, Shaojin Chen, Fu Jin, Shufang Wang, Xiaolan Huang, Licheng Wang, Xiaocui Zhou, Xi Chen, Xiaodai Cui, Jieqiong Li, Jun Tai, Xiong Zhu

## Abstract

CoronaVac (Sinovac), an inactivated vaccine for SARS-CoV-2, has been widely used for immunization. However, analysis of the underlying molecular mechanisms driving CoronaVac-induced immunity is still limited. Here, we applied a systems biology approach to understand the mechanisms behind the adaptive immune response to CoronaVac in a cohort of 50 volunteers immunized with 2 doses of CoronaVac. Vaccination with CoronaVac led to an integrated immune response that included several effector arms of the adaptive immune system including specific IgM/IgG, humoral response and other immune response, as well as the innate immune system as shown by complement activation. Metabolites associated with immunity were also identified implicating the role of metabolites in the humoral response, complement activation and other immune response. Networks associated with the TCA cycle and amino acids metabolic pathways, such as phenylalanine metabolism, phenylalanine, tyrosine and tryptophan biosynthesis, and glycine, serine and threonine metabolism were tightly coupled with immunity. Critically, we constructed a multifactorial response network (MRN) to analyze the underlying interactions and compared the signatures affected by CoronaVac immunization and SARS-CoV-2 infection to further identify immune signatures and related metabolic pathways altered by CoronaVac immunization. These results suggest that protective immunity against SARS-CoV-2 can be achieved via multiple mechanisms and highlights the utility of a systems biology approach in defining molecular correlates of protection to vaccination.

## Introduction

The ongoing coronavirus disease 19 (COVID-19) pandemic, caused by severe acute respiratory syndrome coronavirus 2 (SARS-CoV-2), is an unprecedented global threat leading to high morbidity and mortality worldwide (Wang et al., 2020a). Since the outbreak began, researchers from around the world have been trying to develop vaccines for COVID-19, with more than 44 candidate vaccines in the clinical development stage and another 151 vaccines in preclinical evaluation as of February, 2021(Hodgson et al., 2021). CoronaVac (Sinovac Life Sciences, Beijing, China), an inactivated vaccine against COVID-19 has shown good immunogenicity in mice, rats, and non-human primates (Wu et al., 2021; Zhang et al., 2021)). After preclinical evaluation, CoronaVac, approved by the WHO recently, has been widely used in China and other countries to immunize different populations, including children and adolescents aged 3-17 years old, adults aged 18-59, and adults aged 60 years and older (Mallapaty, 2021)

Although the efficacy of CoronaVac has been assessed in large clinical trials involving thousands of subjects, the underlying molecular processes and cellular mechanisms by which biological messages stimulate the immune response remains poorly understood (Wu et al., 2021; Zhang et al., 2021). Previous analysis of COVID-19 vaccines has mainly focused on evaluating immunogenicity and safety, as well as characterizing immune cell types and/or cytokines (Polack et al., 2020; Wu et al., 2021; Zhang et al., 2021). Protective immunity induced by vaccines not only involves the response of the innate and adaptive immune cells, but also induces profound changes in cellular proteomic and metabolic pathways, increasing the capacity of these immune cells to respond to secondary stimulation. Systems vaccinology, which uses high-throughput cellular and molecular omics technologies, allows the immune response to be comprehensively studied to increase our understanding of vaccine-induced immunity (Li et al., 2017; Nakaya et al., 2015; Tsang et al., 2014). Being able to quickly determine vaccine efficacy, and specific protein and metabolite changes would aid in controlling epidemics and pandemics when speed is a critical factor.

Blood proteomics and metabolomics have provided valuable insights into the early events of vaccine-induced immune response (Best et al., 2018; Camponovo et al., 2020; Rieckmann et al., 2017). For example, proteomic signatures after vaccination have been used to predict vaccine-induced T cell responses in multiple studies, and different classes of vaccines have been shown to induce distinct protein expression patterns (Camponovo et al., 2020). The coordinated action of the immune system induced by vaccines resembles a social network. This enables complex immunological tasks to be performed beyond the sum of the functions of individual immune cells (Li et al., 2017; Rieckmann et al., 2017). Furthermore, increasing evidence have linked trained immunity to epigenetic and metabolic regulation that involve a number of central cellular metabolic pathways such as glycolysis, oxidative phosphorylation, glutaminolysis, as well as fatty acids and cholesterol-synthesis pathways (Arts et al., 2016; Domínguez-Andrés et al., 2019; Voss et al., 2021). Metabolic rewiring is a crucial step for the induction of trained immunity after immunization, but many questions remain including which metabolic pathways are involved (e.g. the role of pentose phosphate pathway or reactive oxygen species metabolism), what immune cells are affected and what specific effects do these metabolic changes have in the affected immune cells (Bekkering et al., 2018). Taken together, proteomics and metabolic studies contribute to the emerging field of systems vaccinology and open up new ways to understand the molecular mechanisms of vaccine-induced immunity.

Beside efficacy, safety evaluation is another important parameter for vaccine evaluation. Recently, a growing body of clinical data suggests that proteomic and metabolic dysregulation are associated with COVID-19 pathogenesis (Shen et al., 2020). For example, acute phase proteins (APPs) including serum amyloid A-1 (SAA1), SAA2, SAA4 and C-reactive protein (CRP) were increased in severe COVID-19 patients, indicating activation of inflammation and the complement system (Shen et al., 2020). This leads to enhanced cytokine and chemokine production, potentially contributing to ‘cytokine storm’, and increases recruitment of macrophages from peripheral blood, which may result in acute lung injury (Chirco and Potempa, 2018). In contrast to infection, the inflammatory response induced by the inactivated vaccine, CoronaVac, should be kept at an appropriate level while still promoting immune cell activation. For this reason, proteomic and metabolomic analysis of vaccine immunized subjects are essential in evaluating the safety of CoronaVac.

To enhance our understanding of the mechanisms behind CoronaVac-induced protection to SARS-CoV-2, we combined multi-omics data, including plasma proteomics, metabolomics, cytokine analysis, and specific IgM/IgG, coupled with computational approaches to construct a global overview of the immune response induced by CoronaVac. The goal of this study is 1) to evaluate the plasma proteomic and metabolomic phenotypes of adaptive immunity to CoronaVac, 2) to delineate the molecular mechanisms that generate protective immunity, and 3) to evaluate the safety of CoronaVac. Understanding how proteins and metabolites affect vaccine immune response has important implications for increasing vaccine efficacy and offering new insights into the molecular mechanisms of protection from SARS-CoV-2 vaccines.

## RESULTS

### Study Design for Integrated Immune Profiling to CoronaVac Vaccination in Humans

Between January and April 2021, fifty participants, aged 18 to 65, were enrolled in Sanya People’s Hospital and immunized with CoronaVac. The detailed descriptions including the sampling date for each participant are shown in **Figure 1A**, and **Table S1**. Participants received two doses of CoronaVac and were vaccinated at 21-33 days (d) intervals. The subjects’ blood samples were collected at baseline (non-injection time point, NJ) prior to vaccination and at about 21 d after the first injection (FJ) and about 14 d after the second injection (SJ) time point, respectively (**Figure 1A**). Throughout the course of this study, we measured SARS-CoV-2-S-specific antibody titers, clinical parameters and cytokines. Plasma proteomics and metabolomics were also analyzed to obtain systems vaccinology data. As a result, each subject was profiled by multiple technologies in a time series. This rich collection of immune profiles, including high-dimensional data from proteomics and metabolomics, provided a unique opportunity to construct an integrated network of the immune response to CoronaVac in humans. For this study, we firstly present each data type separately. Second, the integrative analysis is presented in a framework of a ‘‘multifactorial response network’’ (MRN). Then, we compared the proteomic and metabolomics signatures affected by CoronaVac vaccination and SARS-CoV-2 infection. The safety of CoronaVac was also evaluated using clinical indicators and cytokines combined with proteomics and metabolomics results. Finally, the underling mechanisms and pathways related to CoronaVac-induced immune response were interpreted through a comprehensive analysis.

**Figure 1.**
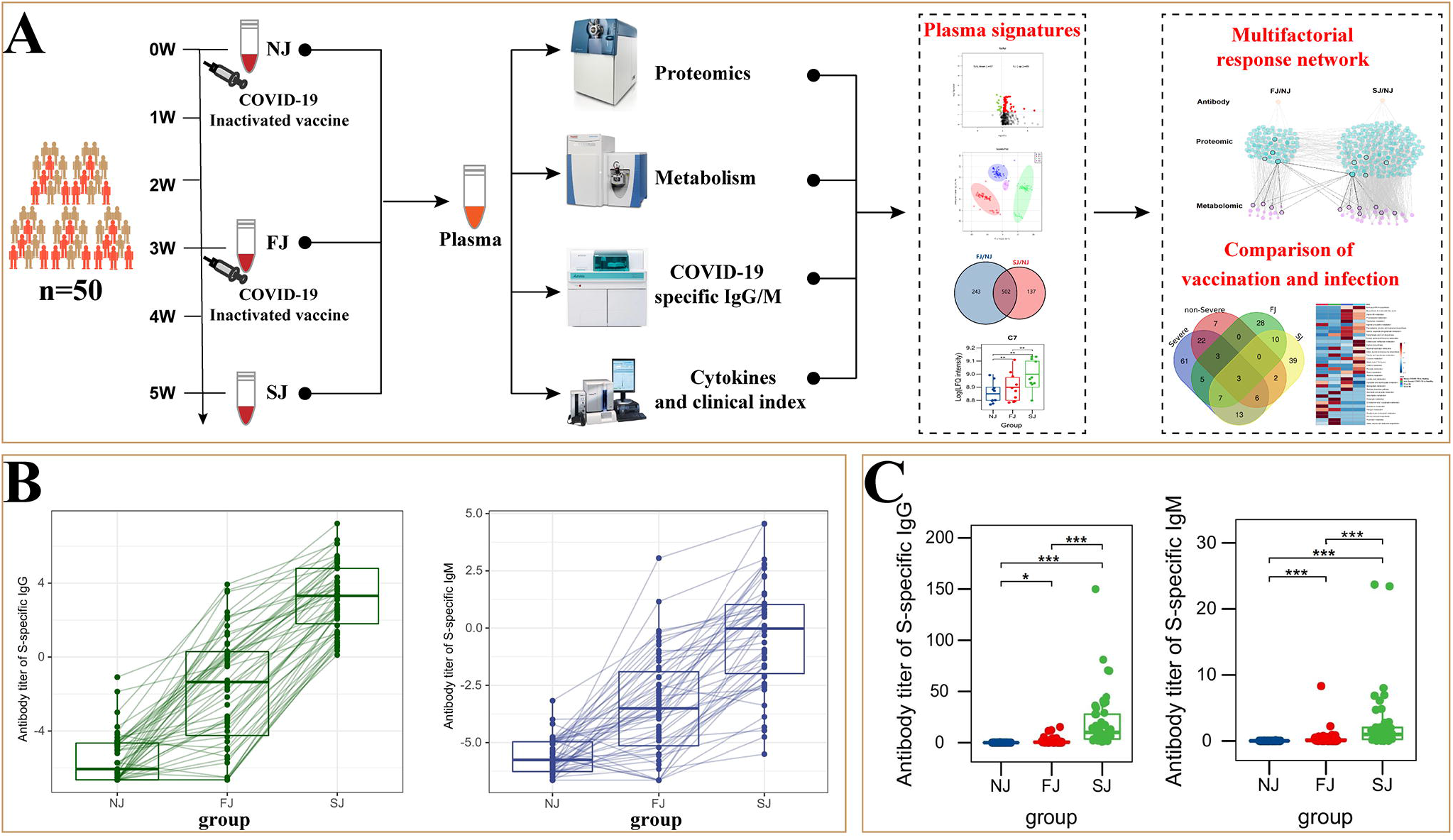
CoronaVac Study Overview and Antibody Expression. (A) Study overview. Fifty subjects were recruited. Different samples were taken at NJ (baseline, day 0), FJ (about 21 days after first immunization) and SJ (about 14 days after second immunization). (B) Representative SARS-CoV-2 S-specific IgG and IgM in each subject. (C) Average levels of SARS-CoV-2 S-specific IgG and IgM in each group. Comparison of response between different time points was done using paired t test. *p < 0.05; **p < 0.01; ***p < 0.001.

### CoronaVac-Induced Antibody Responses

To ensure the effectiveness of vaccination, we assessed the antibody response to SARS-CoV-2 induced by CoronaVac. At baseline, none of the participants had any detectable S-specific IgG and IgM antibodies. The seroconversion rates of IgG were 18 (36%) at FJ point versus 50 (100%) of 50 participants at SJ point, and the seroconversion rates of IgM were 2 (4%) at FJ point versus 25 (50%) of 50 participants at SJ point. The dynamic changes of IgG and IgM to SARS-CoV-2 are shown in **Figure 1B** and illustrates that the IgG antibody levels did not significantly increase until after the second dose of the vaccine. Additionally, as shown in **Figure 1C**, the levels of IgM and IgG were 2.562±4.806 s/co and 19.691±26.86 s/co at SJ point, significantly higher than that at FJ point (IgM 0.388±1.202 s/co, IgG 1.667±3.21 s/co) and the baseline (IgM 0.024±0.017 s/co, IgG 0.039±0.075 s/co). Taken together, these results show that CoronaVac induced antibody responses in all subjects involved in this study. This was consistent with the increased antibody levels reported in clinical trials (Wu et al., 2021; Zhang et al., 2021).

### Plasma Proteomic Signatures after CoronaVac Vaccination

Based on the LC-MS/MS data, we identified 5054 peptides in total with 4400 peptides (87.1%) supported by ≥ 2 MS/MS counts. These peptides were then mapped and 387 proteins (73.7%) were identified with FDR ≤ 1% (**Figure S1A** and **Table S2**). Of the 387 proteins, 116 were identified as differentially expressed proteins (DEPs) in vaccine immunized samples compared to baseline (NJ). The number of DEPs and magnitude of fold change were increased in the SJ group, indicating that the alterations in plasma proteins became more extensive after the second dose of CoronaVac (**Figure S2A-D and Table S3**). As expected, gene ontology (GO) terms for DEPs were highly enriched in processes involved in known immune-related functions, such as complement activation, regulation of complement activation, humoral immune response, and regulation of humoral immune response (**Figure 2A**). Interestingly, besides complement and coagulation cascades, KEGG analysis also identified COVID-19 and pertussis pathways as significantly associated with CoronaVac immunization (**Figure 2B**). GO and KEGG analysis of DEPs from FJ vs NJ and SJ vs NJ also showed similar enrichment (**Figure S2 E-H**). As a COVID-19 inactivated vaccine, CoronaVac induced DEPs naturally enriched to COVID-19 pathway, suggesting that the adaptive immunity induced by CoronaVac were similar with immunity induced by SARS-CoV-2 infection. Similar to our findings, *Reche* et al., (Reche, 2020) also found that pertussis vaccines contain cross-reactive epitopes with SARS-CoV-2, and thus there may a general mechanism for cross-protection between SARS-CoV-2 and pertussis. Cluster analysis showed that the expression of DEPs can be divided into 8 different trends (**Figure S3A**).

**Figure 2.**
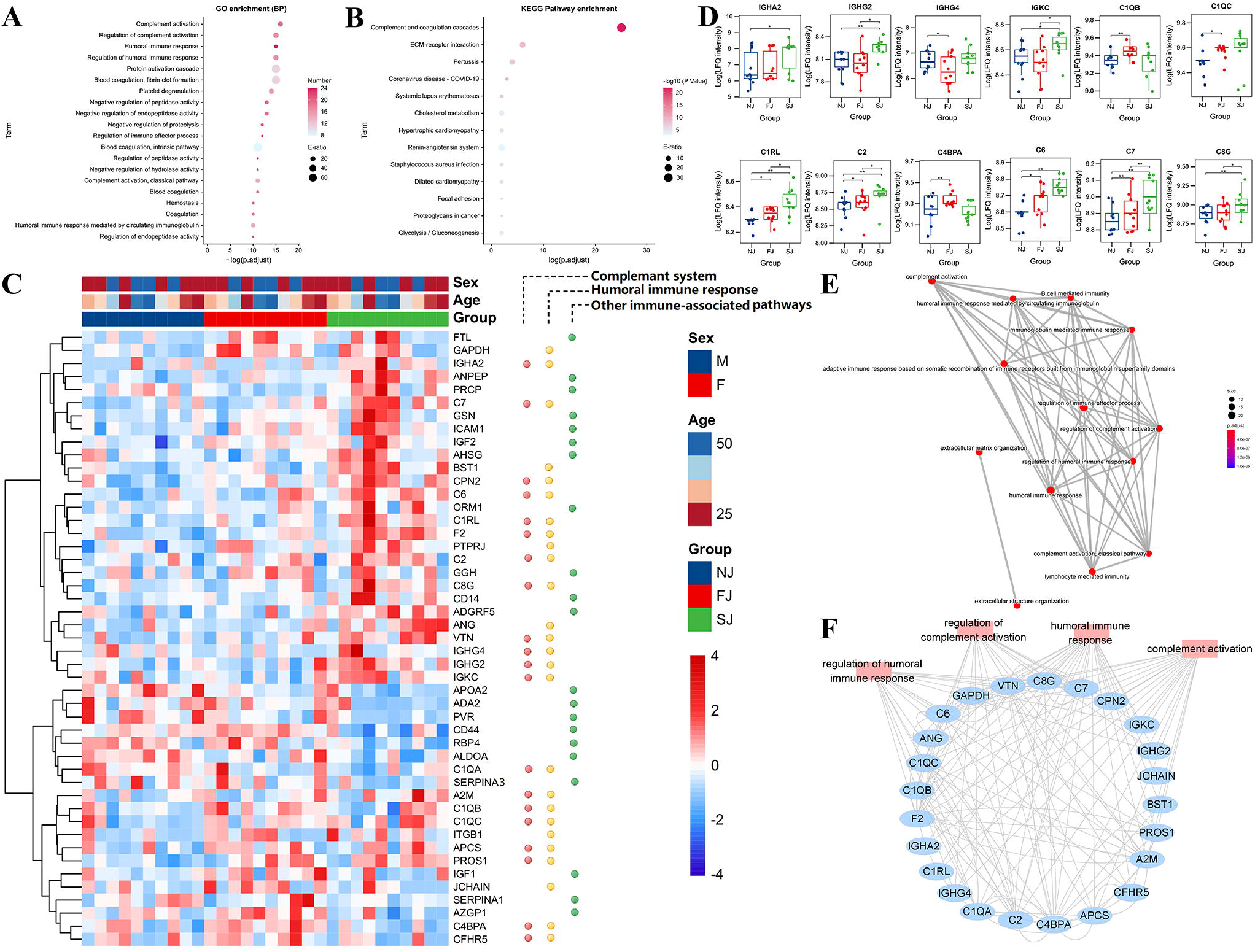
Enrichment and Distribution of Immune Related Proteins. (A) GO analysis of the total DEPs from FJ vs NJ and SJ vs NJ. (B) The KEGG pathway enrichment analysis of the total DEPs from FJ vs NJ and SJ vs NJ. (C) Heatmap of selected proteins from three enriched pathways: complement response, humoral immune response, and other immune associated-pathways. (D) The expression level comparison of humoral immune related proteins with significant differences. Statistical significance was determined by paired two-sided Welch’s t test. *p < 0.05; **p < 0.01; ***p < 0.001. (E) Network of GO modules related to the immune response defined by clusterProfiler v0.1.4. (F) Interaction diagram of proteins involved in the humoral immune response, regulation of humoral immune response, complement activation, and regulation of complement activation.

### Evidence of Humoral and Complement Response Activation After CoronaVac Immunization

An important goal of systems vaccinology is to evaluate the adaptive response and innate immunity to vaccination. Here, proteomics data showed that adaptive immunity, especially humoral and complement response were activated by vaccination. As shown in **Figure 2C**, 47 DEPs belonged to three major pathways: activation of the complement response, humoral immune response, and other immune-associated pathways. At the SJ time point, multiple immunoglobulin heavy chains, including IGHA2, IGHG2, IGHG4, and IGKC were highly upregulated (**Figure 2C** and **Figure 2D**). Consistent with our report, expression of *IGHG2* and *IGKC* were also enhanced in subjects immunized with meningococcal vaccine, and correlated with immunogenicity (O’Connor et al., 2017). IGHG2 encodes the constant region of the heavy chain of IgG2, which is the predominant IgG subclass directed against specific antigens (Calonga-Solís et al., 2019). Increased expression of IgG in the plasma of vaccinated subjects also supports this finding (**Figure 1B**).

Complements were reported to have a protective role in enhancing virus neutralization by antibodies (Kurtovic and Beeson, 2021). It is also a central regulator for adaptive immune responses due to its essential role in delivering co-stimulatory signals via engagement of complement receptors on B and T cells (West et al., 2020). Although several recent studies have implicated complement activity or impairment in severe COVID-19 patients, the potential involvement of complement factors in protective immunity has been largely ignored for SARS-CoV-2 (Shen et al., 2020; Shu et al., 2020; Tian et al., 2020). In this study, we found that several complements, including C1QB, C1QC, C1RL, C2, C4BPA, C6, C7, and C8G were significantly increased at FJ and/or SJ time point (**Figure 2D**). Consistently, complements including C1QB were also found to be upregulated in blood cells early after yellow fever (YF17D) vaccination (Gaucher et al., 2008). These molecules have been shown to have a potential role in inducing dendritic cell maturation (Hosszu et al., 2012). Taken together, this suggests a possible role for the complement system in establishing protective immunity in response to CoronaVac vaccination.

In addition, other immune associated proteins were also observed to be increased in vaccine immunized subjects. For instance, it has been reported that ICAM-1 expression on DCs plays a crucial role in mediating T cell migration and activation (Boyd et al., 1988; Comrie et al., 2015). In this study, the level of ICAM-1 was significantly increased in immunized samples, implying the activation of T cells after vaccination. Human monocyte differentiation antigen CD14 is a pattern recognition receptor that enhances the innate immune response and was significantly increased after the first immunization (Wu et al., 2019). Additionally, SERPINA1, important for the development of neutrophils, was also significantly increased in CoronaVac immunized samples. Similarly, the levels of SERPINA1 were also increased in BCG immunized samples, which indicates the induction of trained immunity in humans by vaccination (Cirovic et al., 2020). Collectively, our results demonstrate the activation of adaptive immunity, especially the humoral and complement responses, after CoronaVac vaccination.

To further understand the functions and interactions of DEPs induced by CoronaVac vaccination, we categorized these proteins based on GO biological processes (GO-BP) using the Cytoscape plug-in ClueGo (**Figure 2E**). All of the modules clustered into two groups, reflecting their functional lineage relationship. Among the 28 resulting functional modules, humoral regulated proteins were enriched for ‘regulation of humoral immune response’, ‘B cell mediated immunity’, ‘humoral immune response’ and ‘immunoglobulin mediated immune response’, and created a complex network. Humoral response was also tightly connected with complement-related modules such as complement activation, regulation of complement activation, and complement activation-classical pathway. Furthermore, the network of lymphocyte-related immunity, containing immunoglobulin proteins, was also significantly connected with the humoral and complement response network. The specialized function of complement, comprised of 12 proteins, was highly enriched for “complement activation” and “regulation of complement activation”. Notably, the proteins belonging to these modules were also connected with each other (**Figure 2F**). Together these results demonstrate that DEPs involved in adaptive immunity formed a complex interactive network which allowed us to further analyze the underlying mechanism of immunity induced by CoronaVac.

### Plasma Metabolomic Signatures Induced by CoronaVac

To ensure reliable results, the PCA score plot, which included the control group, model group and QC samples, is shown in **Figure S1B**. QC samples (purple) clustered tightly together, reflecting the stability of the instrument and showed that the quality of all the LC MS data generated in this study was satisfactory. In addition, during the entire experiment, 88.68%, 37.8%, and 3% of the metabolite relative standard deviation (RSD) in the QC samples were within 30%, 15%, and 5%, respectively (**Figure S1C**). These results demonstrate the reliability of the analytical methods used.

We identified 1190 metabolites (**Table S2**). Compared to the baseline (NJ), 882 differentially expressed metabolites (DEMs), containing amino acids, lipids and other important serum metabolites, were identified. A summary of the number of DEMs between the vaccination and pre-vaccination cohorts is shown in **Figure 3A**. Further details of the DEMs from FJ vs NJ and SJ vs NJ are shown in **Figure S4** and **Table S4**. Not surprisingly, orthogonal partial least squares discrimination analysis (OPLS-DA) showed a high degree of separation between groups, illustrating evident differences in their plasma metabolite profiles (**Figure S1B**). Consistent with our proteomic data, the DEMs were also divided into 8 different clusters (**Figure S3B**). Metabolite Set Enrichment Analysis of DEMs revealed CoronaVac immunization had a significant impact on amino acid metabolism, especially pathways involving valine, leucine and isoleucine biosynthesis (**Figure 3B**). Moreover, the TCA cycle and other amino acid metabolism, such as phenylalanine metabolism, phenylalanine, tyrosine and tryptophan biosynthesis, and glycine, serine and threonine metabolism were also affected by vaccination. In comparison, DEMs of samples from subjects immunized with Zostavax and a *Francisella tularensis* vaccine were enriched for metabolites associated with the TCA cycle and 2-oxocarboxylic acid metabolism (Goll et al., 2020; Li et al., 2017). Notably, these results revealed distinct metabolomic signatures in the immune response to CoronaVac which provided key insights into vaccine-induced antiviral responses.

**Figure 3.**
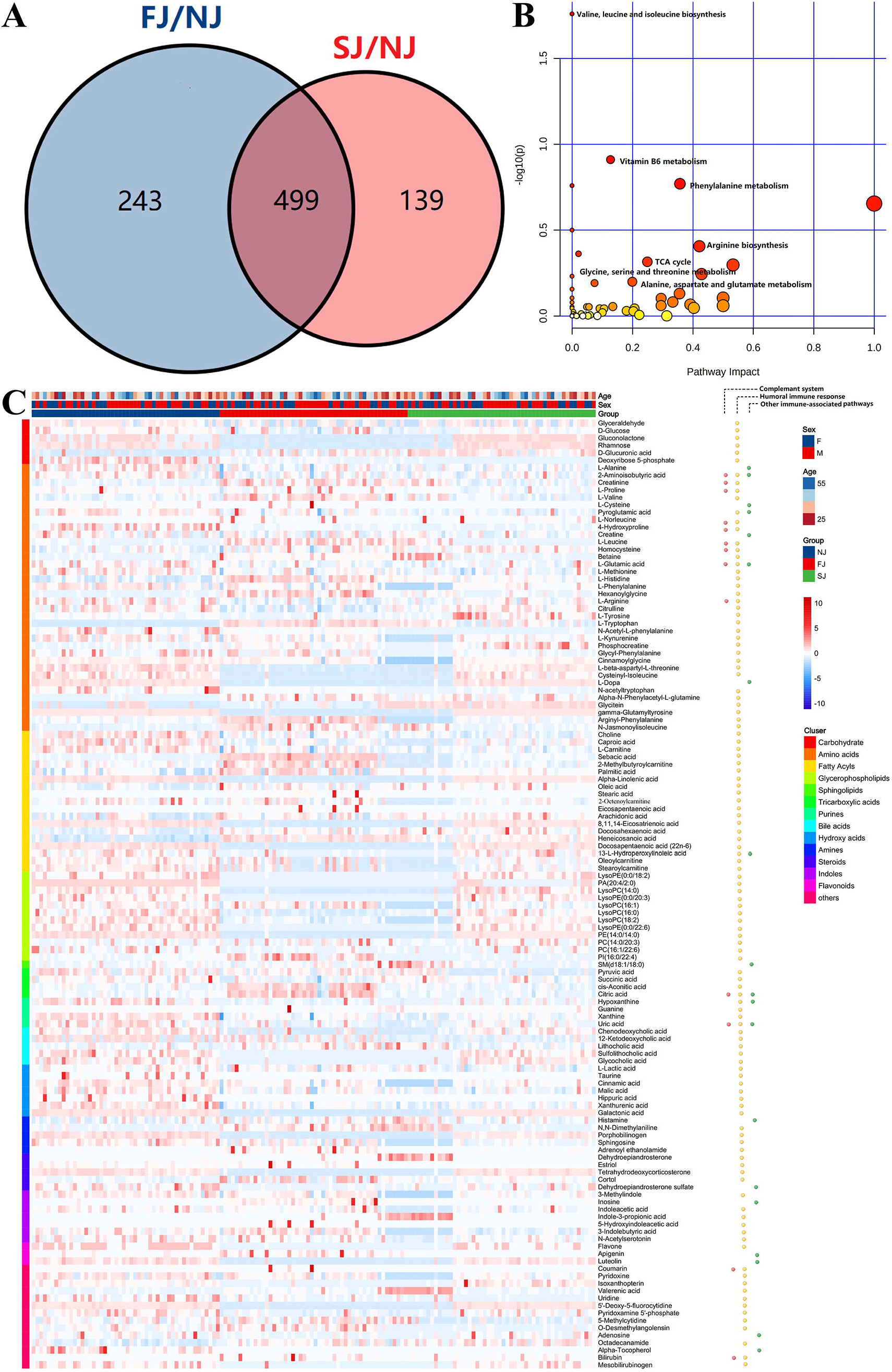
Enrichment and Distribution of Immune Related Metabolites. (A) Venn diagram showing the number of differentially expressed metabolites (DEMs). (B) The KEGG pathway enrichment analysis of the total DEMs from FJ vs NJ and SJ vs NJ. (C) Heatmap of the DEMs associated with three enriched pathways: complement system, humoral immune response, and other immune associated-pathways.

### Metabolites associated with the Humoral and Complement Immune Response are altered after CoronaVac Immunization

A growing body of evidence suggests that metabolic pathways such as the TCA cycle, amino acid metabolism, and lipid metabolism, play an essential role in adaptive and innate immunity. To elucidate the role of metabolites in vaccine immunity, functional metabolites associated with immunity were selected for further analysis (**Figure S5**). Consistent with our proteomic analysis, 128 significant differentially expressed metabolites including carbohydrates, amino acids, and several types of lipids were involved in the three enriched biological processes identified in the proteomic analysis (**Figure 3C**). Production of antibody after vaccination is mainly mediated by plasma cells and requires large quantities of amino acids and glycosylation sugars to properly build and fold antibodies (Lam and Bhattacharya, 2018). In this study, we found significant changes in TCA intermediary metabolites (pyruvate, citrate, succinate, malate, and lactate) in vaccine immunized samples, implying involvement of the TCA cycle in vaccination (**Figure 4A**). The balance between energy and amino acid metabolism is essential for antibody production and in this study, we also identified more than 20 amino acids that were significantly altered in vaccine immunized samples compared to baseline samples (NJ). We found metabolites involved in arginine and proline metabolism were significantly changed after vaccination (**Figure 4B**). Arginine has been reported to participate in antibody synthesis (Fan et al., 2015). Additionally, several amino acids involved in phenylalanine metabolism were also significantly altered in vaccinated samples (**Figure 4C**). Phenylalanine metabolism is associated with humoral autoimmune diseases, which suggests an underlying function in the humoral response (Blackmore et al., 2020). Amino acids and metabolites involved in tryptophan metabolism were also significantly altered after CoronaVac immunization (**Figure S6A-B**). In addition, several fatty acids were also significantly changed in vaccinated samples (**Figure S6C**). These fatty acids may be important for the initial expansion of the endoplasmic reticulum during plasma cell differentiation (Dufort et al., 2014). Collectively, our results show that vaccination changes energy metabolism, amino acid metabolism and fatty acid metabolism. The balances of these metabolic pathways may play an important role in antibody production following vaccination.

**Figure 4.**
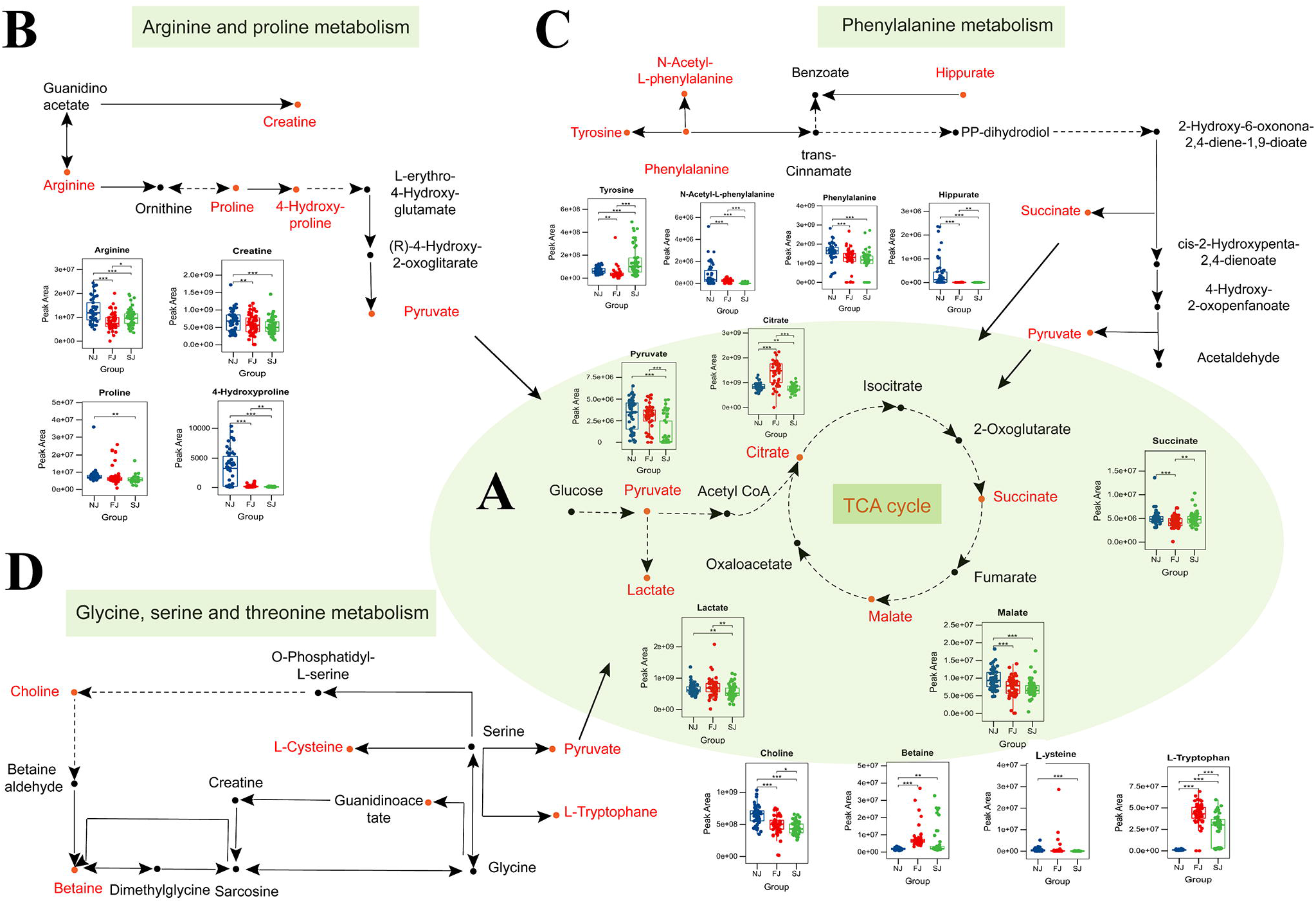
Changes in Amino Acid Levels, Carbohydrate Levels, and Their Metabolic Pathways After Vaccination. (A) Circulating levels of TCA metabolites in plasma. Serum levels of metabolites involved in the TCA cycle were significantly changed when comparing vaccination samples with baseline samples. (B) Significant changes were seen in the levels of some intermediates of the arginine and proline metabolism pathways in plasma of vaccine-immunized samples. (C) Significant changes were observed in the levels of some intermediates of the phenylalanine metabolism pathways in plasma of vaccine-immunized samples. (D) Significant changes were observed in the levels of metabolites involved in glycine, serine and threonine metabolism pathways after vaccination. Statistical significance was determined by paired two-sided Welch’s t test. *p < 0.05; **p < 0.01; ***p < 0.001.

Glycine, serine and threonine metabolism have been reported to affect complement-mediated killing (Cheng et al., 2019). In this study, L-cysteine, betaine, choline, and L-tryptophan, which are enriched in the glycine, serine and threonine metabolism pathway, were significantly changed in vaccine immunized samples (**Figure 4D**). In addition to antibody production and complement response, innate immunity was also linked to metabolic regulation and involved a number of central cellular metabolic pathways such as the TCA cycle, oxidative phosphorylation, as well as fatty acids and cholesterol-synthesis. Our data indicated that the innate immune system was activated after vaccination and this was accompanied by a series of related metabolites changing. For example, several fatty acids that were associated with proteins from the pathway “other immune-associated pathways”, have also been reported to be associated with a pro-inflammatory macrophage phenotype (**Figure 3C**) (Oishi et al., 2017). Taken together, our data suggests that altered metabolites may be a crucial step for adaptive immunity, and further integrated analysis will increase understanding of the underlying mechanism for how CoronaVac induces protection.

### Multifactorial Response Network (MRN) Reveals a Connection between Antibody Response, Proteins, and Metabolites Following CoronaVac Immunization

To integrate antibody, proteomics, and metabolomics data, we constructed a MRN for mechanistic analysis. The MRN had a dense network of 329 nodes and 1395 connections. This hierarchical structure allows an overview of the super-network and reveals the complex relationship of the adaptive immune response induced by CoronaVac (**Figure 5A**).

**Figure 5.**
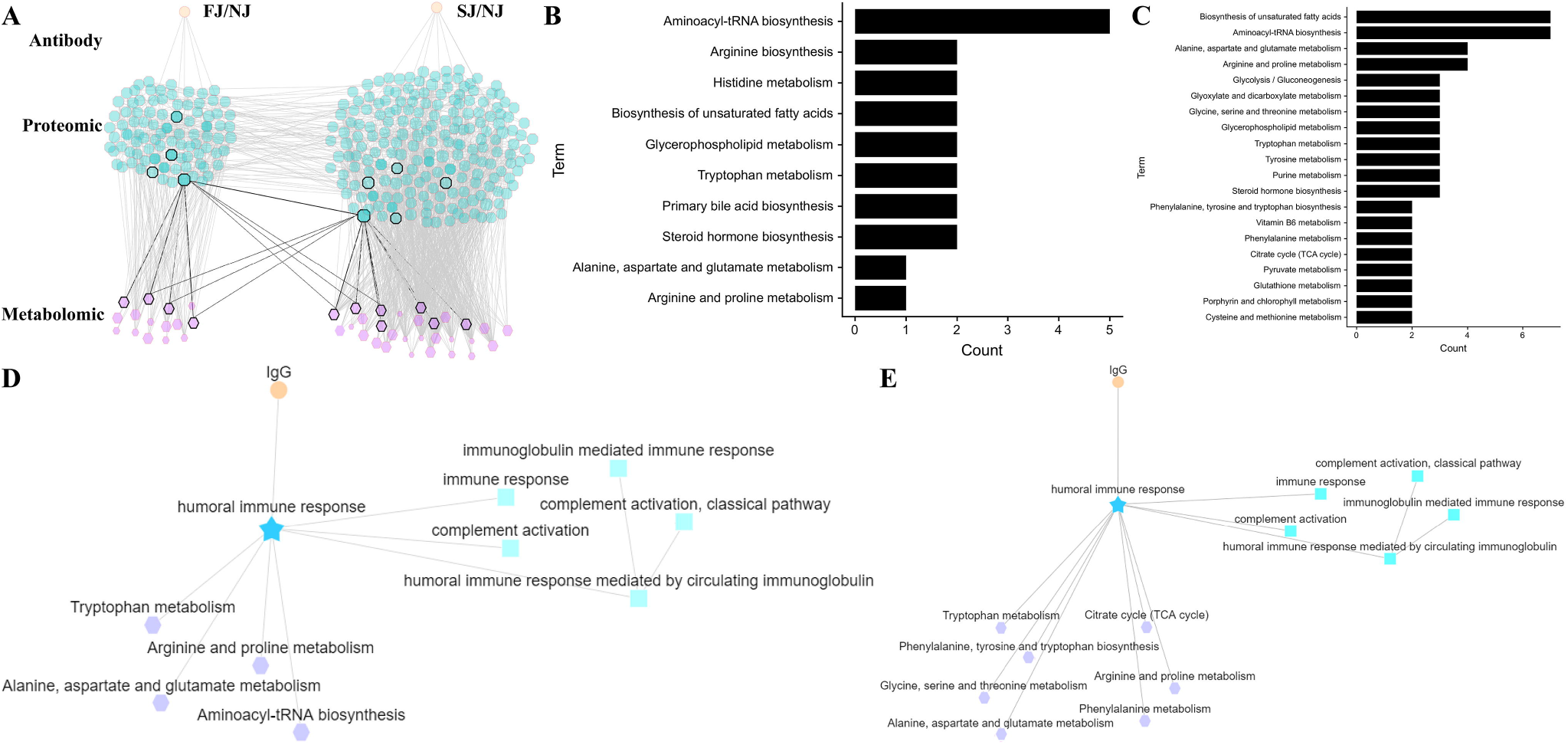
MRN analysis of Metabolomics, Proteomics, and Antibodies. (A) MRN consists of correlation networks using data from antibody, proteomics, and metabolomics. Each node is a child network of one data type. The links between nodes were established by weight. (B) The top pathways for metabolite networks correlated with IgG levels at FJ time point. (C) The top pathways for metabolite networks correlated with IgG levels at SJ time point. (D) Connections between IgG, humoral response associated network and metabolite networks at FJ time point are shown. (E) Connections between IgG, humoral response associated network and metabolite networks at SJ time point are shown.

A humoral response-related network was obtained to further investigate the underlying mechanisms following vaccination. As described above, levels of IGHA2, IGHG2, IGHG4, and IGKC correlated with antibody titers and were significantly increased after vaccination (**Figure 2D**). These proteins that were enriched in the humoral immune response were also connected with other immune response pathways such as immunoglobulin mediated immune response, humoral immune response mediated by circulating immune and complement activation (**Figure S5D**). They were also correlated with the levels of IgG and expression of many enriched metabolites involved in biosynthesis of unsaturated fatty acids, the TCA cycle and several other amino acids pathways (**Figure S5D**). This suggests a connection between the antibody response and these metabolic pathways (**Figure 5B** and **Figure 5C**). We hypothesize that aminoacyl-tRNA biosynthesis, biosynthesis of unsaturated fatty acids and amino acid metabolic pathways such as tryptophan metabolism, alanine, aspartate and glutamate metabolism, and arginine and proline metabolism, which were all significantly altered following CoronaVac immunization at FJ and SJ timepoint are involved in and affects the antibody response (**Figure 5D**). In addition, after the second immunization, the TCA cycle, phenylalanine metabolism and the phenylalanine, tyrosine and tryptophan biosynthesis pathways were further altered by vaccination and may also be associated with the antibody response (**Figure 5E**). Finally, the humoral immune response pathway was also connected with complement activation and complement activation-classical pathway (which also showed tight correlation with the metabolism pathways mentioned above). Taken together, these data suggest that high activity in these metabolic pathways discussed above was detrimental to the humoral immune response induced by CoronaVac and they combined a complex network in which many proteins and metabolites are involved.

### Comparison of the Proteomic and Metabolomic Signatures Induced by CoronaVac Immunization and SARS-CoV-2 Infection

To gain an insight into the mechanisms underlying the responses to vaccines against SARS-CoV-2, we combined data from a published paper (Shen et al., 2020) to compare differences in the proteomic and metabolomic signatures affected by vaccination and SARS-CoV-2 infection. Notably, DEPs in vaccination and infection were different (**Figure 6A**). Interestingly, GO terms of DEPs after vaccination were highly enriched in processes involved in known immune-related functions such as complement activation, regulation of complement activation, humoral immune response, and regulation of humoral immune response (**Figure 2A**). In comparison, DEPs from COVID-19 patients were mainly enriched in platelet degranulation, regulation of hemostasis, blood coagulation, humoral immune response, complement activation, and acute-phase response (**Figure 6B** and **Figure 6C**) **(Shen et al., 2020)**. Surprisingly however, several pathways related to immunity including humoral immune response, complement activation, regulation of complement activation and regulation of humoral immune response, which were induced by CoronaVac vaccination, were also strongly enriched in severe COVID-19 patients (**Figure 6D**). Enriched pathways unique to CoronaVac vaccination were complement activation-classical pathway and humoral immune response mediated by circulating immunoglobulin (**Figure 6E**). It was reported that SARS-CoV-2 infection induced dysregulation of macrophages, platelet degranulation and complement system pathways (Shen et al., 2020). Thus, DEPs related to macrophage function, platelet degranulation, and immune response were selected for further expression analysis (**Figure 6F**). Compared to infection, many circulating immunoglobulins and complements were elevated in CoronaVac immunized samples. For example, IGHA2 and IGHG2 were significantly increased after vaccination while they were not significant changed after infection. Both vaccines and infections lead to the activation of the complement system, but the phenotypes and underlying functions may be different. As shown in **Figure 6F**, C1QB, C1QC, and C7 were significantly increased after vaccination, while C5 and C8A were increased after infection. In addition, SARS-CoV-2 infection induced elevated acute phase proteins, including SAA1, SAA2, SAA4, CRP, alpha-1-antichymotrypsin (SERPINA3), and serum amyloid p-component (SAP/APCS) (Shen et al., 2020). As expected, except for SERPINA3 most acute phase proteins showed no changes following CoronaVac immunization. The potential function of SERPINA3 in COVID-19 should be studied further as it may be involved in immune cell infiltration (Xia et al., 2021). Besides acute phase proteins, a low platelet count was also reported to be associated with severe COVID-19 and mortality (Lippi et al., 2020). CoronaVac immunized samples did not alter the expression of plasma proteins such as platelet-expressing chemokines proplatelet basic protein (PPBP), platelet factor 4 (PF4) and other 10 platelet associated proteins. Interestingly, proteins such as APOH, CLEC3B, HSPA5, and AHSG also showed opposite trends between COVID-19 patients and CoronaVac immunized subjects (Shen et al., 2020). These results indicate that CoronaVac does not suppress platelet function.

**Figure 6.**
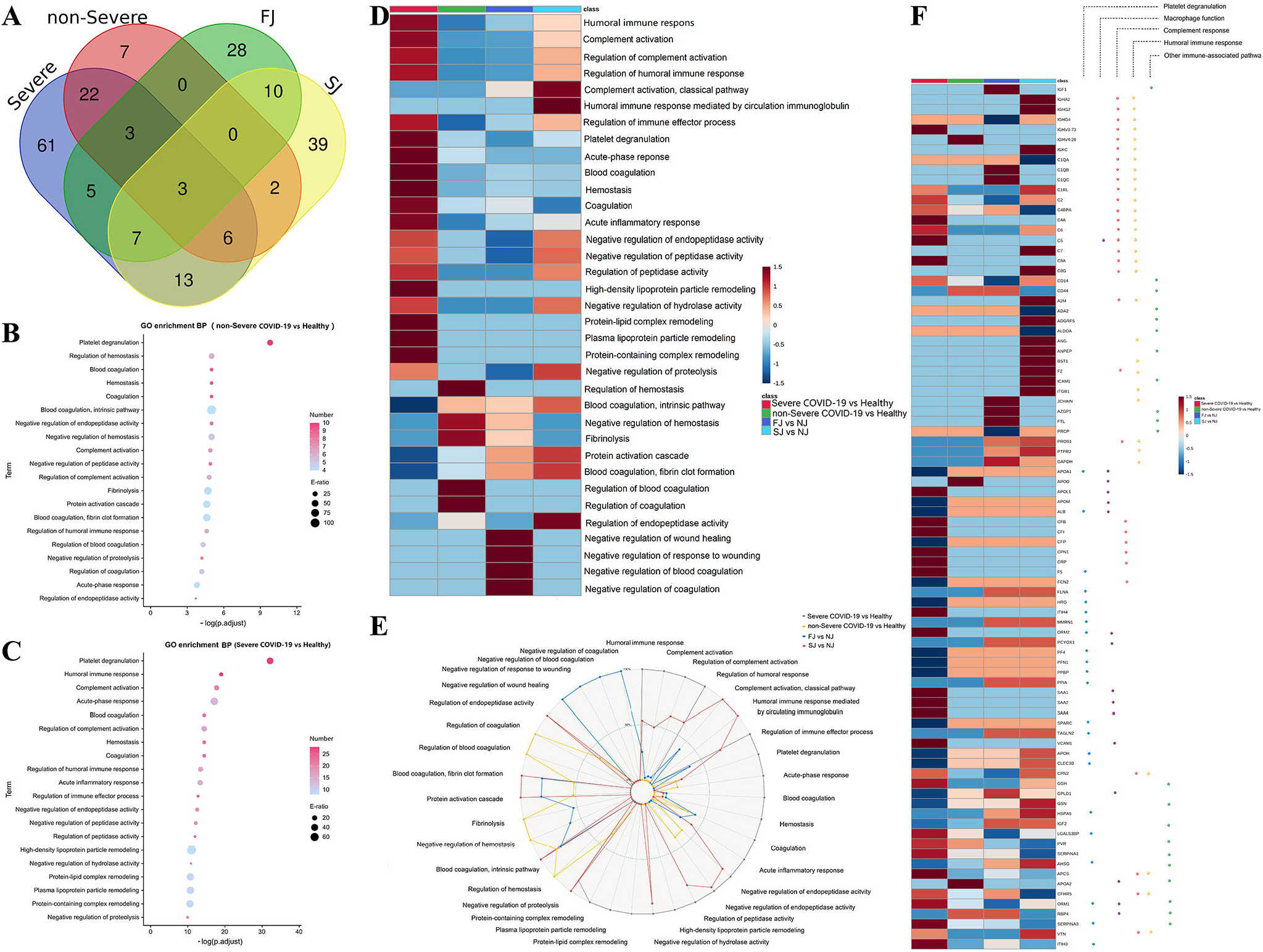
Comparison of DEPs and their enriched pathways in severe COVID-19 infection, non-severe COVID-19 infection, FJ and SJ group. (A) Venn diagram showing the number of differentially expressed proteins in severe COVID-19, non-severe COVID-19, FJ and SJ. (B) GO analysis of DEPs from severe COVID-19 vs healthy. (C) KEGG analysis of DEPs from non-severe COVID-19 vs healthy. (D) Heatmap of the top 20 GO terms enriched from DEPs in severe COVID-19 vs healthy, non-severe COVID-19 vs healthy, FJ vs NJ and SJ vs NJ. (E) Radar map of the top 20 GO terms enriched from DEPs in severe COVID-19 vs healthy, non-severe COVID-19 vs healthy, FJ vs NJ and SJ vs NJ, respectively. -Log10 p values of GO terms were used to make the radar map. (F) Heatmap of DEPs from vaccination and infection that were related to platelet degranulation macrophage function, complement response, humoral immune response andother immune-associated pathway.

Metabolomics phenotypes induced by vaccination and infection were also significantly different (**Figure 7A**). Interestingly, there were some common differentially expressed metabolites identified between vaccination and infection, however, the expression trends were almost the opposite. For example, the levels of tryptophan, leucine, betaine, isoleucine, citrate, and valine were decreased in COVID-19 patients but increased in vaccination samples. Conversely, guanosine, kynurenine and uracil were increased in COVID-19 patients but decreased in vaccination samples (**Figure 7B**). Similar to vaccination, the metabolomics data for COVID-19 infection also revealed a significant impact on amino acid metabolism. However, the types of amino acid metabolism pathways affected by vaccination and infection were different: COVID-19 infection mainly affected pathways involved in valine, leucine and isoleucine biosynthesis, aminoacyl-tRNA biosynthesis, and arginine biosynthesis (**Figure 7C** and **Figure 7D**). Compared to infection, vaccination with CoronaVac significantly altered other pathways including: vitamin B6 metabolism, biosynthesis of unsaturated fatty acids, phenylalanine metabolism, tryptophan metabolism, arginine and proline metabolism, and glycine, serine and threonine metabolism (**Figure 7E** and **Figure 7F**). These data suggest that high activity in amino acid metabolism, such as phenylalanine metabolism, tryptophan metabolism, arginine and proline metabolism, and glycine, serine and threonine metabolism as well as fatty acids pathway might be detrimental to humoral immune responses. Altogether, vaccination caused a variety of unique proteomic and metabolomic changes compared to infection, and these differences may play an essential role in the protective mechanism induced by CoronaVac.

**Figure 7.**
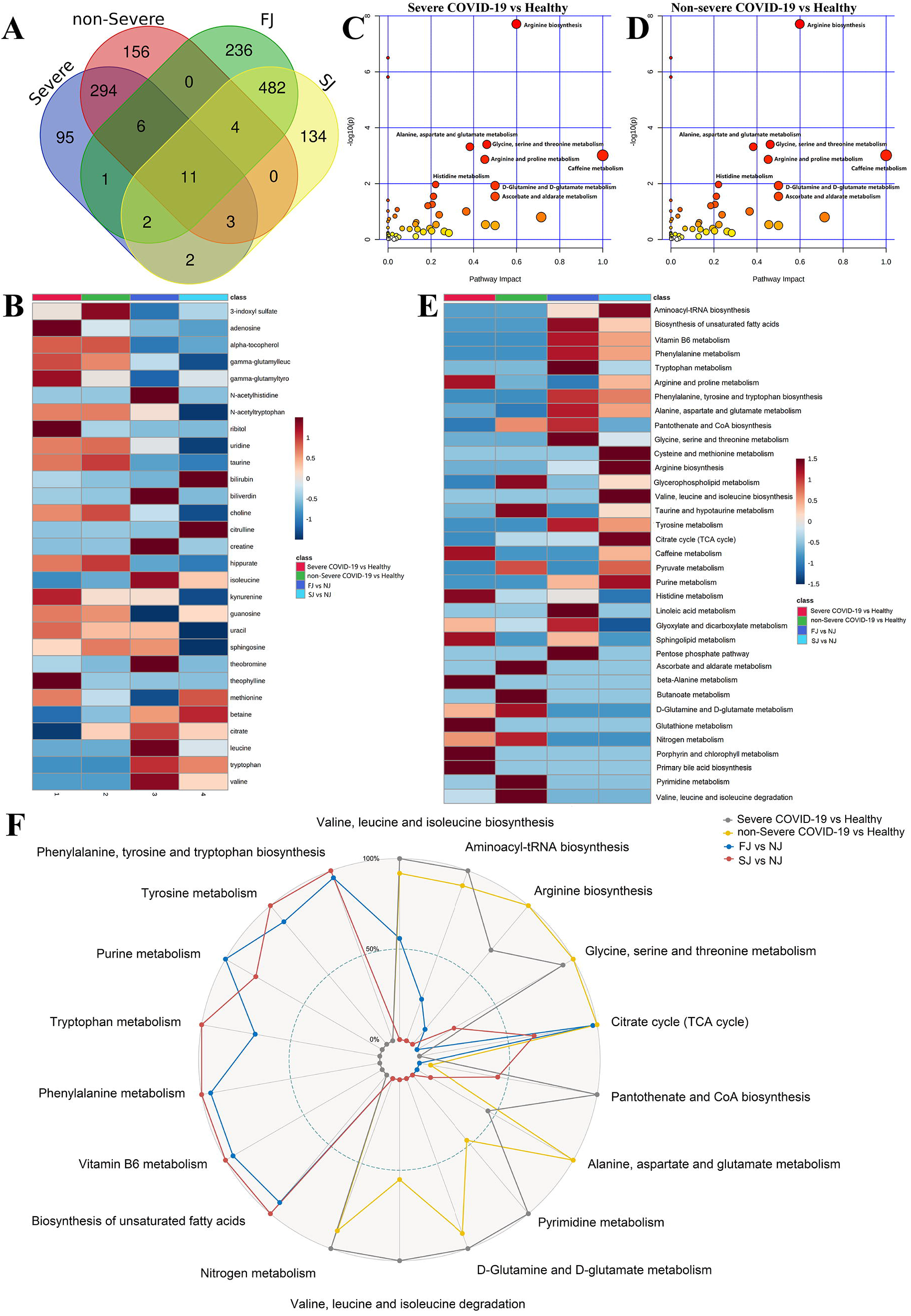
Comparison of DEMs and their enriched pathways in severe COVID-19 infection, non-severe COVID-19 infection, FJ and SJ group. (A) Venn diagram showing the number of differentially expressed metabolites in severe COVID-19, non-severe COVID-19, FJ and SJ. (B) KEGG analysis of DEMs from severe COVID-19 vs healthy. (C) KEGG analysis of DEMs from non-severe COVID-19 vs healthy. (D) Heatmap of the top 20 KEGG terms enriched from DEMs in severe COVID-19 vs healthy, non-severe COVID-19 vs healthy, FJ vs NJ and SJ vs NJ. (E) Radar map of the top 20 KEGG terms enriched from DEMs in severe COVID-19 vs healthy, non-severe COVID-19 vs healthy, FJ vs NJ and SJ vs NJ, respectively. -Log10 p values of GO terms were used to make the radar map. (F) Heatmap of the common metabolites shared between CoronaVac immunization and infection.

### Safety Evaluation of CoronaVac

Safety evaluation is also an important goal in systems vaccinology. Previous data have shown that SARS-CoV-2 infection causes a variety of disorders including “cytokine storm”, excessive inflammation, and suppressed platelet degranulation. COVID-19 may activate the ability of T cells to stimulate pro-inflammatory cytokines. Clinical reports show that both mild and severe forms of disease result in cytokine secretion, particularly IL-6, IL-1β, TNF, GM-CSF, and IL-1α (Wang et al., 2020b). In this study, no significant differences in inflammatory cytokines were observed in immunized samples; this was consistent with previous studies which also reported a lack of upregulated cytokines for subjects immunized with CoronaVac (**Table S5** and **Figure S7**) (Wu et al., 2021; Zhang et al., 2021)

Clinical laboratory-related factors are also associated with disease severity in COVID-19 patients. It was observed that SARS-CoV-2 induced higher expression of lymphocyte (Lym), monocyte (Mon), and white blood cells (WBC). A low platelet count was also reported to be associated with severe COVID-19 disease and mortality (Lippi et al., 2020). In this study, we analyzed 22 clinical measurements, including count and proportion of blood cells [white blood cells (WBC), red blood cell (RBC), neutrophils (Neu), lymphocyte (Lym), eosinophils (Eos), monocytes (Mon), basophils (Bas)], hemoglobin-related clinical indicators and platelet-related clinical indicators to evaluation the safety of CoronaVac. Compared to baseline (NJ), no significant changes except for eosinophils were observed in plasma from the immunized group (**Table S6** and **Figure S8**). Furthermore, the proteomic data also demonstrated that most acute phase proteins were not significantly altered during CoronaVac immunization (**Figure 2B**). Altogether, our data suggests that CoronaVac does not cause serious adverse reactions in immunized subjects.

## Discussion

This study is, to our knowledge, the first study to combine multi-omics data, including plasma proteomics and metabolomics with cytokines, clinical index, and specific IgM/IgG for system biology analysis of CoronaVac. Previously, systems biology have been applied to identify signatures of immune responses to vaccination and have provided insights into the mechanisms of immune responses induced by different vaccines such as the live attenuated yellow fever vaccine (YF-17D) (Gaucher et al., 2008; Querec et al., 2009), smallpox vaccine (Reif et al., 2009), malaria vaccine (Kazmin et al., 2017) and influenza vaccines (Nakaya et al., 2015). These insights can be used to guide novel strategies for vaccine evaluation and design.

It was observed that CoronaVac can induce quick antibody responses which may be suitable for emergency use and is of vital importance during the COVID-19 pandemic (Wu et al., 2021; Zhang et al., 2021). However, the underlying molecular mechanisms induced by CoronaVac is still a mystery. Here, we combined proteomics and metabolomics to identify the signatures associated with the immune response to CoronaVac as changes in genes, proteins and metabolites may reflect the immune status. It may also reveal the underlying molecular mechanisms involved in vaccine-induced immunity. Our results showed that IgG and other immunoglobulin components such as IGHA2, IGHG2, IGHG4, IGKC, and IGKV2-24, were significantly elevated in vaccine immunized samples. IGHA2, IGHG2 and IGHG4 are encoded by the immunoglobulin heavy chain (IGH) constant genes (*IGHC*), while IGKC and IGKV2-24 are encoded by the subgroup of immunoglobulin light chain genes (Walther et al., 2015). These results demonstrate the activation of B-cell and plasma cells after vaccination which play an essential role in antiviral immunity. In addition, the high expression of IGHG2 and IGHG4 might demonstrate that IgG2 and IgG4 are the main type of IgG induced by CoronaVac. Previous multifactorial models have also revealed integrated networks on humoral immunity induced by vaccination against various other diseases (Camponovo et al., 2020; Li et al., 2014; Li et al., 2017; O’Connor et al., 2017). For instance, herpes zoster (HZ) induced changes in genes and metabolites associated with adaptive immunity and showed that sterol metabolism was tightly coupled with immunity (Li et al., 2017). In Li et al., (Li et al., 2014) gene expression data was collected from five different vaccine cohorts and revealed distinct transcriptional signatures for antibody responses to different types of vaccines. The study suggested that gene expression predictors for antibody response are probably not ‘universal’ but are dependent on the type of vaccine, which is consistent with the proposal that different types of vaccines would induce similar signatures of immunogenicity. Consistent with other reports, our findings also support the humoral response as a key factor for antiviral immunity and that the immunity induced by CoronaVac showed distinctive characteristics.

Metabolites associated with humoral immunity were also revealed to play a potential role in the underlying mechanism for CoronaVac-induced immunity in this study. In our data, the essential intermediary metabolites in the TCA cycle were changed in vaccine immunized samples. These TCA cycle intermediary metabolites play a crucial role in regulating the immune system with some activating the immune response and others suppressing it (Choi et al., 2021). In cells with infection or other stresses, TCA cycle intermediates may accumulate and regulate inflammatory gene expression (Tannahill et al., 2013; Williams and O’Neill, 2018). Surprisingly, we found that most of the TCA cycle intermediates including pyruvate, citrate, malate and lactate were significantly decreased after vaccination and were negatively associated with IgG. These results suggest that vaccination does not trigger a severe inflammatory response like infection does, and that balancing TCA cycle intermediates could impact the immune response. Furthermore, the balance between energy and amino acid metabolism is essential for antibody production. As the central link to energy metabolism, metabolites in the TCA cycle are also involved in different amino acid metabolic pathways. In this study, vaccination also caused changes in amino acids and amino acid metabolic pathways such as arginine and proline metabolism, phenylalanine metabolism and glycine, serine and threonine metabolism. Although most of the metabolites were reported to be associated with antibody production, whether these amino acid metabolites also have other immunological roles after vaccination requires further investigation (Blackmore et al., 2020; Cheng et al., 2019; Fan et al., 2015).

Complements can also be activated by antigen-specific antibodies and therefore, can contribute to the adaptive immune responses (Mellors et al., 2020). However, few studies have focused on vaccine-induced complement responses. There have been several reports on the role of complement in COVID-19 disease in humans, however all were focused on innate complement activation that occurs during acute infection(Shen et al., 2020; Tian et al., 2020). These studies have generally concluded that excess complement activity can contribute to severe disease pathology. Importantly, the potential involvement of complement factors in protective immunity has been largely ignored for SARS-CoV-2 but has been defined for other viruses, bacteria, and protozoa. Here, our results highlight that several kinds of complements were significantly increased after vaccination, implying that it might play a protective role and are consistent with Kurtovic’s viewpoint (Kurtovic and Beeson, 2021). In other diseases, subjects immunized with the yellow fever vaccine, YF-17D, also showed activation of the complement system. C3a, a product of the classical complement enzymatic pathway was increased at day 7 after immunization (Querec et al., 2009). In addition, immunity to many viral and nonviral pathogens relies on antibodies and antibody-mediated neutralization. This has been demonstrated *in vitro* for the human pathogens; West Nile virus (Mehlhop et al., 2009), Nipah virus (Johnson et al., 2011), and others (Mellors J et al., 2020) using a combination of human and non-human antibodies and complements. It has also been suggested that COVID-19 patients with mild disease generally report normal serum concentrations of complement proteins, which suggests that these immune mediators may be able to contribute to immunity and reduce disease severity (Du et al., 2021). In line with this, an examination of >6000 COVID-19 patients found that individuals with a dysregulated complement system were more prone to developing severe disease than those with a healthy complement system (Ramlall et al., 2020). Furthermore, distinct components of the complement pathway were found to be essential for activating the innate immune response, including IFN-stimulated responsive element and nuclear factor-κB reporters, against viral infection (Wang et al., 2012). Therefore, for most individuals, complement activation might contribute to reduced disease severity, whereas for a smaller percentage of individuals, the complement system might be dysregulated and associated with increased susceptibility to severe disease. The implications for antibody-complement interactions in virus neutralization and immunity should be further investigated and may have important implications for antibody-based vaccination strategies against SARS-CoV-2.

According to a phase 1/2 clinical trial, no vaccine-related serious adverse events were reported (Wu et al., 2021; Zhang et al., 2021). Similarly, no “cytokine storm” and changes in clinical index, except for slightly elevated eosinophils were observed after vaccination in this study. In addition, systems vaccinology has also been applied for safety evaluation. Mizukami et al. (Mizukami et al., 2014) applied a systems vaccinology approach in a rat model to predict safety and batch-to-batch consistency of influenza vaccines. In this study, most acute phase proteins, such as SAA1, SAA2, SAA4, CRP, SERPINA3, and SAP/APCS, which were increased in severe COVID-19 patients were not changed in our CoronaVac immunized samples. The plasma proteomic and metabolic signatures of vaccine immunized samples were also different from that of COVID-19 patients, further supporting the idea that this inactivated vaccine was safe.

It should be noted that although secreted proteins and metabolites can directly reflect the immune status, they contain less information than the transcriptome and will be analyzed in future studies. Additionally, this was a single-center prospective study with a relatively small sample size and missing values, which are common in LC-MS data and may affect data interpretation in studies with smaller sample sizes. Therefore, future large-sized cohort studies are warranted to confirm the findings in this study.

In conclusion, our systems vaccinology study showed that CoronaVac immunization was safe and induced humoral immune responses against SARS-CoV-2. These results support the approval for emergency use of CoronaVac in China. The differentially expressed proteins and metabolites identified formed a complex network that resulted in vaccine-induced antiviral immunity. MRN analysis and comparison between CoronaVac vaccination and SARS-CoV-2 infection indicated that humoral and complement responses as well as several metabolic pathways, including the TCA cycle, phenylalanine metabolism, tryptophan metabolism, arginine, proline metabolism and fatty acids pathways, might be essential for protective immunity induced by CoronaVac.

## Material and methods

### Experimental design and participant recruitment

A total of 50 subjects immunized with the COVID-19 vaccine, CoronaVac, were recruited. Written informed consent was obtained from each subject and protocols were approved by Institutional Review Boards of Sayan People’s Hospital.

### COVID-19-specific IgM/IgG ELISA

The S-specific IgG and IgM were detected using a chemiluminescence quantitative kit (Auto Biotechnology, Zhengzhou, China). Plates were coated with either SARS-CoV-2 recombinant antigens or mouse anti-human IgM monoclonal antibody. Ten μL of sample, 20 μL of microparticle solution and 100 μL of sample diluent were mixed and incubated for 37 min at 37□. The plates were then washed and enzyme. Plates were washed and conjugates were added, and incubated for 17 min at 37□ Plates were washed and chemiluminescence developed using 50 μL Chemiluminescent Substrate A and 50 μL of Chemiluminescent Substrate B. The antibody titer was measured using the AutoLumo A2000 Plus. Results with S/CO≥1.00 were considered positive while S/CO<1.00 were considered negative.

### Evaluation of clinical characteristics and markers

Complete information, including count and proportion of blood cells [white blood cells (WBC), red blood cell (RBC), neutrophils (Neu), lymphocyte (Lym), eosinophils (Eos), monocytes (Mon), basophils (Bas)], hemoglobin-related clinical indicators including hemoglobin(HGB), hematocrit (HCT), mean corpuscular volume (MCV), mean corpuscular hemoglobin (MCH), mean corpuscular hemoglobin concentration (MCHC), red blood cell distribution width-coefficient of variation (RDW-CV), red blood cell distribution width-standard deviation (RDW-SD), and platelet-related clinical indicators including platelet (PLT), platelet volume distribution width (PDW), plateletcrit (PCT) were analyzed using the Sysmex XE-2100 (Sysmex Corporation).

### Multiplex cytokine assays

Concentrations of granulocyte-macrophage colony stimulating factor (GM-CSF), interferon (IFN)-γ, Interleukin (IL)-1β, IL-12, IL-13, IL-18, IL-2, IL-4, IL-5, IL-6 and tumor necrosis factor (TNF)-α in plasma were determined using a bead-based, 11-plex Th1/Th2 human ProcartaPlex immunoassay (Thermo Fisher Scientific) according to the manufacturer’s instructions. Fluorescence was measured with a Luminex 200 system (Luminex Corporation) and analyzed with ProcartaPlex Analyst 1.0 software (Thermo Fisher Scientific). Only cytokines above the limit of detection were included for further analysis.

### Plasma proteomics

Ten of the fifty subjects were randomly selected for plasma proteomic analysis. To remove highly abundant interfering proteins in human plasma, a multiple-affinity removal system liquid chromatography (LC) column (High Select™ Top14 Abundant Protein Depletion Mini Spin Columns; Thermo Fisher Technologies, Santa Clara, CA, USA) was used. Briefly, plasma samples loaded onto a multiple-affinity removal system LC column were eluted into fractions which contained low-abundance proteins while highly abundant proteins were removed. The eluted product was used for MS analysis. The concentration of plasma proteins was measured and 50 µg protein samples were prepared for mass spectrometry analysis.

Plasma from each sample was lysed in 100 μL lysis buffer (8 M urea in 100 mM triethylammonium bicarbonate, TEAB) at 25◦ C for 30 min. The lysates were reduced using 5 mM Tris (2-carboxyethyl) phosphine (Pierce, Rockford, IL, USA) and incubated at 37°C for 30 min with shaking (300 rpm). For alkylation, 15 mM Iodoacetamide (Sigma-Aldrich, St. Louis, MO, USA) was added to each sample and incubated at 25°C, with agitation at 300 rpm for 1 h in the dark. Proteins were trypsin digested overnight at 37°C. Mass spectrometry-grade trypsin gold (Promega, Madison, WI, USA) was used with an enzyme-to-protein ratio of 1:50. The dried peptides were L loading buffer (1% formic acid, FA; 1%acetonitrile, ACN). Ten μL of sample was used for LC-MS/MS analysis on an Orbitrap Fusion Lumos in data dependent acquisition (DDA) mode coupled with Ultimate 3000 (Thermo Fisher Scientific, Waltham, MA, USA). The samples were loaded and separated by a C18 trap column (3mm 0.10×20mm), packed with C18 reverse phase particle (1.9mm 0.15×120mm, Phenomenex, Torrance, California, USA). The peptides were eluted using a 75 min nonlinear gradient: 7% B for 11 min, 15–25% B for 37 min, 25–40% B for 20 min, 40–100% B for 1 min, 100% B for 6min (Buffer A, 0.1% FA in ddH2O; Buffer B, 0.1% FA and 80% ACN in ddH2O; flow rate, ∼600 nL/min). All reagents used were MS grade.

The parameters for MS detection were as follows: full MS survey scans were performed in the ultra-high-field Orbitrap analyzer at a resolution of 120,000 and trap size of 500,000 ions over a mass range from 300 to 1400 m/z. MS/MS scan were detected in IonTrap and the 20 most intense peptide ions with charge states 2 to 7 were subjected to fragmentation via higher energy collision-induced dissociation (5×10^3^ AGC target, 35 ms maximum ion time).The resultant mass spectrometry data were analyzed using Maxquant (Version 1.6.17) and the protein search database used was the *Homo sapiens* FASTA database downloaded from UniprotKB (UP000005640.fasta). The following search parameters were used for Maxquant: precursor ion mass tolerance was set at 20 ppm; full cleavage by trypsin was selected; a maximum of two missed cleavages was allowed; static modifications were set to carbamidomethylation (+57.021464) of cysteine, and variable modifications were set to oxidation (+15.994915) of methionine and acetylation (+42.010565) of peptides’ N-termini. The remaining parameters followed the default Maxquant setup. For protein identification, the following criteria was used: (1) peptide length ≥6 amino acids; (2) FDR ≤1% at the PSM, peptide and protein levels. Peptides were quantified using the peak area derived from their MS1 intensity. The intensity of unique and razor peptides was used to calculate the protein intensity.

### Plasma metabolomics

Participant plasma, which were immediately stored at -80°C upon collection, was thawed on ice. To ensure data quality for metabolic profiling, pooled quality control samples were prepared by mixing equal amounts of plasma (0.75 mL) from 150 samples. The pretreatment of the QC samples was performed in parallel and was the same as the study samples. The QC samples were evenly inserted between each set of runs to monitor the stability of the large-scale analysis. Plasma samples were extracted by adding 400 μL of MeOH/ACN (1:1, v/v) solvent mixture to 100 μL of plasma (2:2:1 ratio, no H_2_O added). The mixtures were shaken vigorously for 5 min and incubated for 1 h at -20°C. Samples were then centrifuged for 10 minutes at 13,500 x g at 4°C and the supernatant was transferred to a new centrifuge tube. To ensure that the metabolites detected were reliable, three platforms were used for shotgun metabolomics. Each supernatant was divided into three fractions: two for reverse-phase/ultra-performance liquid chromatography (RP/UPLC)-MS/MS methods with positive ion-mode electrospray ionization (ESI) and negative-ion mode ESI, and one for hydrophilic interaction liquid chromatography (HILIC)/UPLC-MS/MS with positive-ion mode ESI.

All UPLC-MS/MS methods used the ACQUITY 2D UPLC system (Waters, Milford, MA, USA) and Q-Exactive Quadrupole-Orbitrap (Thermo Fisher Scientific™, San Jose, USA) and TripleTOF 5600+ (AB SCIEX, MA, USA) with ESI source and mass analyzer. In the UPLC-MS/MS method, the QE was operated under positive electron spray ionization (ESI) coupled with a C18 column (UPLC BEH C18, 2.1 × 100 mm, 1.7 μm; Waters). The mobile solutions used in the gradient elution were water and methanol containing 0.1% FA. When the QE was operated under negative ESI mode, the UPLC method used a C18 column eluted with mobile solutions containing methanol and water in 6.5 mM ammonium bicarbonate at pH 8. The UPLC column used in the hydrophilic interaction method was a HILIC column (UPLC BEH Amide, 2.1 × 150 mm, 1.7 μm; Waters), and the mobile solutions consisted of water and acetonitrile with 9 mM ammonium formate at pH 8.0; the TripleTOF 5600+ was operated under positive ESI mode. The mass spectrometry analysis alternated between MS and data-dependent MS2 scans using dynamic exclusion. The scan range was 70-1,000 m/z. After raw data pre-processing, peak finding/alignment, and peak annotation using MSDIAL software, metabolite identifications were supported by matching the retention time, accurate mass, and MS/MS fragmentation data to MSDIAL software database and online MS/MS libraries (Human Metabolome Database (HMDB, https://hmdb.ca). Open database sources including KEGG and MetaboAnalyst, Human Metabolome Database, were used to identify metabolic pathways.

### Statistical analysis

All missing values were substituted with 1/5 the minimal value. Missing values were imputed with the minimal value for each feature. The influence of age and sex to the proteomic profiling was assessed using partial least squares regression. For each comparison, the log2 fold-change (log2 FC) was calculated by averaging the paired fold change for each participant. A two-sided paired Welch’s t test was also performed. Statistical significance and differentially expressed proteins were assigned as p value <0.05 and absolute log2 FC >0.25. The correlation between IgG and metabolites was analyzed using Pearson’s correlation (p < 0.05).

Orthogonal partial least squares discrimination analysis (OPLS-DA) and partial least squares-discriminate analysis (PLS-DA) was conducted using MetaboAnalyst 5.0 (http://www.metaboanalyst.ca/MetaboAnalyst/). Volcano plots were calculated using a combination of fold-change and paired Welch’s t test. The intensity data of these regions were used in box-plot analysis and hierarchical cluster analysis. Heat maps of differential metabolites and relationships were displayed using the Multi Experiment Viewer software (MeV, version 4.7.4). Pathway analysis and visualization were performed using the Metaboanalyst5.0 web portal (http://www.metaboanalyst.ca/).

Connected networks of the differentially expressed proteins were built and analyzed in BINGO. Metscape was used to build the network of metabolites, analyze the correlation of these different metabolites and visualize the networks. Heat map, column chart, radar map, cluster were made using R packets.

## Data Availability

All data have been presented in the manuscript.

## Ethical approval

This study was approved by the Ethics Committee of the Sanya People’s Hospital (SYPH-2021-26).

## Data sharing

No additional data available.

## Transparency declaration

The lead author and guarantor affirms that the manuscript is an honest, accurate, and transparent account of the study being reported; that no important aspects of the study have been omitted; and that any discrepancies from the study as planned and registered have been explained.

## Acknowledgments

We thank all the participants. We gratefully acknowledge the participation of Fan-Xing Biological Technology Co., Ltd. (Tianjin) for the support of bioinformatics analysis with their Analysis Platform, and thanks Miss. Yan Li for her contribution.

## References

Arts, R.J., Novakovic, B., Ter Horst, R., Carvalho, A., Bekkering, S., Lachmandas, E., Rodrigues, F., Silvestre, R., Cheng, S.C., Wang, S.Y., et al. (2016). Glutaminolysis and Fumarate Accumulation Integrate Immunometabolic and Epigenetic Programs in Trained Immunity. Cell metabolism 24, 807–819.

Bekkering, S., Arts, R.J.W., Novakovic, B., Kourtzelis, I., van der Heijden, C., Li, Y., Popa, C.D., Ter Horst, R., van Tuijl, J., Netea-Maier, R.T., et al. (2018). Metabolic Induction of Trained Immunity through the Mevalonate Pathway. Cell 172, 135–146.e139.

Best, S.A., De Souza, D.P., Kersbergen, A., Policheni, A.N., Dayalan, S., Tull, D., Rathi, V., Gray, D.H., Ritchie, M.E., McConville, M.J., and Sutherland, K.D. (2018). Synergy between the KEAP1/NRF2 and PI3K Pathways Drives Non-Small-Cell Lung Cancer with an Altered Immune Microenvironment. Cell metabolism 27, 935–943.e934.

Blackmore, D., Li, L., Wang, N., Maksymowych, W., Yacyshyn, E., and Siddiqi, Z.A. (2020). Metabolomic profile overlap in prototypical autoimmune humoral disease: a comparison of myasthenia gravis and rheumatoid arthritis. Metabolomics : Official journal of the Metabolomic Society 16, 10.

Boyd, A.W., Wawryk, S.O., Burns, G.F., and Fecondo, J.V. (1988). Intercellular adhesion molecule 1 (ICAM-1) has a central role in cell-cell contact-mediated immune mechanisms. Proceedings of the National Academy of Sciences of the United States of America 85, 3095–3099.

Calonga-Solís, V., Malheiros, D., Beltrame, M.H., Vargas, L.B., Dourado, R.M., Issler, H.C., Wassem, R., Petzl-Erler, M.L., and Augusto, D.G. (2019). Unveiling the Diversity of Immunoglobulin Heavy Constant Gamma (IGHG) Gene Segments in Brazilian Populations Reveals 28 Novel Alleles and Evidence of Gene Conversion and Natural Selection. Frontiers in immunology 10, 1161.

Camponovo, F., Campo, J.J., Le, T.Q., Oberai, A., Hung, C., Pablo, J.V., Teng, A.A., Liang, X., Sim, B.K.L., Jongo, S., et al. (2020). Proteome-wide analysis of a malaria vaccine study reveals personalized humoral immune profiles in Tanzanian adults. eLife 9.

Cheng, Z.X., Guo, C., Chen, Z.G., Yang, T.C., Zhang, J.Y., Wang, J., Zhu, J.X., Li, D., Zhang, T.T., Li, H., et al. (2019). Glycine, serine and threonine metabolism confounds efficacy of complement-mediated killing. Nature communications 10, 3325.

Chirco, K.R., and Potempa, L.A. (2018). C-Reactive Protein As a Mediator of Complement Activation and Inflammatory Signaling in Age-Related Macular Degeneration. Frontiers in immunology 9, 539.

Choi, I., Son, H., and Baek, J.H. (2021). Tricarboxylic Acid (TCA) Cycle Intermediates: Regulators of Immune Responses. Life (Basel, Switzerland) 11.

Cirovic, B., de Bree, L.C.J., Groh, L., Blok, B.A., Chan, J., van der Velden, W., Bremmers, M.E.J., van Crevel, R., Händler, K., Picelli, S., et al. (2020). BCG Vaccination in Humans Elicits Trained Immunity via the Hematopoietic Progenitor Compartment. Cell host & microbe 28, 322–334.e325.

Comrie, W.A., Li, S., Boyle, S., and Burkhardt, J.K. (2015). The dendritic cell cytoskeleton promotes T cell adhesion and activation by constraining ICAM-1 mobility. The Journal of cell biology 208, 457–473.

Domínguez-Andrés, J., Joosten, L.A., and Netea, M.G. (2019). Induction of innate immune memory: the role of cellular metabolism. Current opinion in immunology 56, 10–16.

Du, H., Dong, X., Zhang, J.J., Cao, Y.Y., Akdis, M., Huang, P.Q., Chen, H.W., Li, Y., Liu, G.H., Akdis, C.A., et al. (2021). Clinical characteristics of 182 pediatric COVID-19 patients with different severities and allergic status. Allergy 76, 510–532.

Dufort, F.J., Gumina, M.R., Ta, N.L., Tao, Y., Heyse, S.A., Scott, D.A., Richardson, A.D., Seyfried, T.N., and Chiles, T.C. (2014). Glucose-dependent de novo lipogenesis in B lymphocytes: a requirement for atp-citrate lyase in lipopolysaccharide-induced differentiation. The Journal of biological chemistry 289, 7011–7024.

Fan, Y., Jimenez Del Val, I., Müller, C., Wagtberg Sen, J., Rasmussen, S.K., Kontoravdi, C., Weilguny, D., and Andersen, M.R. (2015). Amino acid and glucose metabolism in fed-batch CHO cell culture affects antibody production and glycosylation. Biotechnology and bioengineering 112, 521–535.

Gaucher, D., Therrien, R., Kettaf, N., Angermann, B.R., Boucher, G., Filali-Mouhim, A., Moser, J.M., Mehta, R.S., Drake, D.R., 3rd, Castro, E., et al. (2008). Yellow fever vaccine induces integrated multilineage and polyfunctional immune responses. The Journal of experimental medicine 205, 3119–3131.

Goll, J.B., Li, S., Edwards, J.L., Bosinger, S.E., Jensen, T.L., Wang, Y., Hooper, W.F., Gelber, C.E., Sanders, K.L., Anderson, E.J., et al. (2020). Transcriptomic and Metabolic Responses to a Live-Attenuated Francisella tularensis Vaccine. Vaccines 8.

Hodgson, S.H., Mansatta, K., Mallett, G., Harris, V., Emary, K.R.W., and Pollard, A.J. (2021). What defines an efficacious COVID-19 vaccine? A review of the challenges assessing the clinical efficacy of vaccines against SARS-CoV-2. The Lancet. Infectious diseases 21, e26–e35.

Hosszu, K.K., Valentino, A., Vinayagasundaram, U., Vinayagasundaram, R., Joyce, M.G., Ji, Y., Peerschke, E.I., and Ghebrehiwet, B. (2012). DC-SIGN, C1q, and gC1qR form a trimolecular receptor complex on the surface of monocyte-derived immature dendritic cells. Blood 120, 1228–1236.

Johnson, J.B., Aguilar, H.C., Lee, B., and Parks, G.D. (2011). Interactions of human complement with virus particles containing the Nipah virus glycoproteins. Journal of virology 85, 5940–5948.

Kazmin, D., Nakaya, H.I., Lee, E.K., Johnson, M.J., van der Most, R., van den Berg, R.A., Ballou, W.R., Jongert, E., Wille-Reece, U., Ockenhouse, C., et al. (2017). Systems analysis of protective immune responses to RTS,S malaria vaccination in humans. Proceedings of the National Academy of Sciences of the United States of America 114, 2425–2430.

Kurtovic, L., and Beeson, J.G. (2021). Complement Factors in COVID-19 Therapeutics and Vaccines. Trends in immunology 42, 94–103.

Lam, W.Y., and Bhattacharya, D. (2018). Metabolic Links between Plasma Cell Survival, Secretion, and Stress. Trends in immunology 39, 19–27.

Li, S., Rouphael, N., Duraisingham, S., Romero-Steiner, S., Presnell, S., Davis, C., Schmidt, D.S., Johnson, S.E., Milton, A., Rajam, G., et al. (2014). Molecular signatures of antibody responses derived from a systems biology study of five human vaccines. Nature immunology 15, 195–204.

Li, S., Sullivan, N.L., Rouphael, N., Yu, T., Banton, S., Maddur, M.S., McCausland, M., Chiu, C., Canniff, J., Dubey, S., et al. (2017). Metabolic Phenotypes of Response to Vaccination in Humans. Cell 169, 862–877.e817.

Lippi, G., Plebani, M., and Henry, B.M. (2020). Thrombocytopenia is associated with severe coronavirus disease 2019 (COVID-19) infections: A meta-analysis. Clinica chimica acta; international journal of clinical chemistry 506, 145–148.

Mallapaty, S. (2021). WHO approval of Chinese CoronaVac COVID vaccine will be crucial to curbing pandemic. Nature 594, 161–162.

Mehlhop, E., Nelson, S., Jost, C.A., Gorlatov, S., Johnson, S., Fremont, D.H., Diamond, M.S., and Pierson, T.C. (2009). Complement protein C1q reduces the stoichiometric threshold for antibody-mediated neutralization of West Nile virus. Cell host & microbe 6, 381–391.

Mellors, J., Tipton, T., Longet, S., and Carroll, M. (2020). Viral Evasion of the Complement System and Its Importance for Vaccines and Therapeutics. Frontiers in immunology 11, 1450.

Mizukami, T., Momose, H., Kuramitsu, M., Takizawa, K., Araki, K., Furuhata, K., Ishii, K.J., Hamaguchi, I., and Yamaguchi, K. (2014). System vaccinology for the evaluation of influenza vaccine safety by multiplex gene detection of novel biomarkers in a preclinical study and batch release test. PloS one 9, e101835.

Nakaya, H.I., Hagan, T., Duraisingham, S.S., Lee, E.K., Kwissa, M., Rouphael, N., Frasca, D., Gersten, M., Mehta, A.K., Gaujoux, R., et al. (2015). Systems Analysis of Immunity to Influenza Vaccination across Multiple Years and in Diverse Populations Reveals Shared Molecular Signatures. Immunity 43, 1186–1198.

O’Connor, D., Clutterbuck, E.A., Thompson, A.J., Snape, M.D., Ramasamy, M.N., Kelly, D.F., and Pollard, A.J. (2017). High-dimensional assessment of B-cell responses to quadrivalent meningococcal conjugate and plain polysaccharide vaccine. Genome medicine 9, 11.

Oishi, Y., Spann, N.J., Link, V.M., Muse, E.D., Strid, T., Edillor, C., Kolar, M.J., Matsuzaka, T., Hayakawa, S., Tao, J., et al. (2017). SREBP1 Contributes to Resolution of Pro-inflammatory TLR4 Signaling by Reprogramming Fatty Acid Metabolism. Cell metabolism 25, 412–427.

Polack, F.P., Thomas, S.J., Kitchin, N., Absalon, J., Gurtman, A., Lockhart, S., Perez, J.L., Pérez Marc, G., Moreira, E.D., Zerbini, C., et al. (2020). Safety and Efficacy of the BNT162b2 mRNA Covid-19 Vaccine. The New England journal of medicine 383, 2603–2615.

Querec, T.D., Akondy, R.S., Lee, E.K., Cao, W., Nakaya, H.I., Teuwen, D., Pirani, A., Gernert, K., Deng, J., Marzolf, B., et al. (2009). Systems biology approach predicts immunogenicity of the yellow fever vaccine in humans. Nature immunology 10, 116–125.

Ramlall, V., Thangaraj, P.M., Meydan, C., Foox, J., Butler, D., Kim, J., May, B., De Freitas, J.K., Glicksberg, B.S., Mason, C.E., et al. (2020). Immune complement and coagulation dysfunction in adverse outcomes of SARS-CoV-2 infection. Nature medicine 26, 1609–1615.

Reche, P.A. (2020). Potential Cross-Reactive Immunity to SARS-CoV-2 From Common Human Pathogens and Vaccines. Frontiers in immunology 11, 586984.

Reif, D.M., Motsinger-Reif, A.A., McKinney, B.A., Rock, M.T., Crowe, J.E., Jr., and Moore, J.H. (2009). Integrated analysis of genetic and proteomic data identifies biomarkers associated with adverse events following smallpox vaccination. Genes and immunity 10, 112–119.

Rieckmann, J.C., Geiger, R., Hornburg, D., Wolf, T., Kveler, K., Jarrossay, D., Sallusto, F., Shen-Orr, S.S., Lanzavecchia, A., Mann, M., and Meissner, F. (2017). Social network architecture of human immune cells unveiled by quantitative proteomics. Nature immunology 18, 583–593.

Shen, B., Yi, X., Sun, Y., Bi, X., Du, J., Zhang, C., Quan, S., Zhang, F., Sun, R., Qian, L., et al. (2020). Proteomic and Metabolomic Characterization of COVID-19 Patient Sera. Cell 182, 59–72.e15.

Shu, T., Ning, W., Wu, D., Xu, J., Han, Q., Huang, M., Zou, X., Yang, Q., Yuan, Y., Bie, Y., et al. (2020). Plasma Proteomics Identify Biomarkers and Pathogenesis of COVID-19. Immunity 53, 1108–1122.e1105.

Tannahill, G.M., Curtis, A.M., Adamik, J., Palsson-McDermott, E.M., McGettrick, A.F., Goel, G., Frezza, C., Bernard, N.J., Kelly, B., Foley, N.H., et al. (2013). Succinate is an inflammatory signal that induces IL-1β through HIF-1α. Nature 496, 238–242.

Tian, W., Zhang, N., Jin, R., Feng, Y., Wang, S., Gao, S., Gao, R., Wu, G., Tian, D., Tan, W., et al. (2020). Immune suppression in the early stage of COVID-19 disease. Nature communications 11, 5859.

Tsang, J.S., Schwartzberg, P.L., Kotliarov, Y., Biancotto, A., Xie, Z., Germain, R.N., Wang, E., Olnes, M.J., Narayanan, M., Golding, H., et al. (2014). Global analyses of human immune variation reveal baseline predictors of postvaccination responses. Cell 157, 499–513.

Voss, K., Hong, H.S., Bader, J.E., Sugiura, A., Lyssiotis, C.A., and Rathmell, J.C. (2021). A guide to interrogating immunometabolism. Nature reviews. Immunology.

Walther, S., Rusitzka, T.V., Diesterbeck, U.S., and Czerny, C.P. (2015). Equine immunoglobulins and organization of immunoglobulin genes. Developmental and comparative immunology 53, 303–319.

Wang, D., Hu, B., Hu, C., Zhu, F., Liu, X., Zhang, J., Wang, B., Xiang, H., Cheng, Z., Xiong, Y., et al. (2020a). Clinical Characteristics of 138 Hospitalized Patients With 2019 Novel Coronavirus-Infected Pneumonia in Wuhan, China. Jama 323, 1061–1069.

Wang, J., Jiang, M., Chen, X., and Montaner, L.J. (2020b). Cytokine storm and leukocyte changes in mild versus severe SARS-CoV-2 infection: Review of 3939 COVID-19 patients in China and emerging pathogenesis and therapy concepts. Journal of leukocyte biology 108, 17–41.

Wang, Y., Tong, X., Zhang, J., and Ye, X. (2012). The complement C1qA enhances retinoic acid-inducible gene-I-mediated immune signalling. Immunology 136, 78–85.

West, E.E., Kunz, N., and Kemper, C. (2020). Complement and human T cell metabolism: Location, location, location. Immunological reviews 295, 68–81.

Williams, N.C., and O’Neill, L.A.J. (2018). A Role for the Krebs Cycle Intermediate Citrate in Metabolic Reprogramming in Innate Immunity and Inflammation. Frontiers in immunology 9, 141.

Wu, Z., Hu, Y., Xu, M., Chen, Z., Yang, W., Jiang, Z., Li, M., Jin, H., Cui, G., Chen, P., et al. (2021). Safety, tolerability, and immunogenicity of an inactivated SARS-CoV-2 vaccine (CoronaVac) in healthy adults aged 60 years and older: a randomised, double-blind, placebo-controlled, phase 1/2 clinical trial. The Lancet. Infectious diseases 21, 803–812.

Wu, Z., Zhang, Z., Lei, Z., and Lei, P. (2019). CD14: Biology and role in the pathogenesis of disease. Cytokine & growth factor reviews 48, 24–31.

Xia, X., Wang, M., Li, J., Chen, Q., Jin, H., Liang, X., and Wang, L. (2021). Identification of potential genes associated with immune cell infiltration in atherosclerosis. Mathematical biosciences and engineering : MBE 18, 2230–2242.

Zhang, Y., Zeng, G., Pan, H., Li, C., Hu, Y., Chu, K., Han, W., Chen, Z., Tang, R., Yin, W., et al. (2021). Safety, tolerability, and immunogenicity of an inactivated SARS-CoV-2 vaccine in healthy adults aged 18-59 years: a randomised, double-blind, placebo-controlled, phase 1/2 clinical trial. The Lancet. Infectious diseases 21, 181–192.

